# Repeat expansions in Parkinson’s disease and parkinsonism across ancestries: insights from a global genetic cohort

**DOI:** 10.64898/2026.06.12.26354932

**Authors:** Lara M. Lange, Catalina Cerquera-Cleves, Ai Huey Tan, Shen-Yang Lim, Njideka U. Okubadejo, Chin-Hsien Lin, Pin-Shiuan Chen, Jung Hwan Shin, Azlina Ahmad-Annuar, Laurel A. Screven, Viorica Chelban, Allison A. Dilliot, André Fienemann, Kamalini Ghosh Galvelis, Henry Houlden, Hirotaka Iwaki, Zane Jaunmuktane, Patrick W. Cullinane, Thomas Warner, Johanna Junker, Yuliia Kanana, Ignacio J. Keller Sarmiento, Christine Klein, Pin-Jui Kung, Hampton L. Leonard, Niccoló E Mencacci, Mike A. Nalls, Raquel Real, Samia Ben Sassi, Joanne Trinh, Dan Vitale, Ana Westenberger, Lesley Y. Wu, Andrew B. Singleton, Huw R. Morris, Katja Lohmann, Cornelis Blauwendraat, Peter Heutink, Zih-Hua Fang, Global Parkinson’s Genetics Program (GP2)

## Abstract

Expanded short tandem repeats contribute to a broad spectrum of neurodegenerative diseases, yet their roles in Parkinson’s disease (PD) and parkinsonism remain incompletely characterized, especially across diverse ancestries. We analyzed short-read whole-genome (WGS) and clinical exome sequencing (CES) data from 38,365 individuals (28,861 WGS; 9,504 CES), encompassing 23,242 patients with PD, 4,729 patients with atypical parkinsonism and 10,394 healthy controls from 11 genetic ancestries. To determine carrier frequencies and characterize repeat structures across diverse ancestries, we genotyped 12 established pathogenic loci where normal, intermediate, and pathogenic alleles can be reliably differentiated using short-read sequencing data. Additionally, we conducted threshold-based associations to determine the minimum threshold associated with increased PD risk in 15,995 individuals (8,591 PD, 7,404 controls) of European ancestry.

Pathogenic repeat expansions were detected in 62 patients (56 PD and 6 atypical parkinsonism) and 5 controls across seven loci (*AR*, *ATXN1*, *ATXN2*, *ATXN3*, *CACNA1A*, *HTT* and *THAP11*), spanning seven ancestries. Among these, *ATXN2* expansions were the most frequently observed in PD and were present in African, East Asian, European and Middle Eastern ancestries. Additionally, intermediate *ATXN2* repeat expansions exhibited a strong, length-dependent association with PD risk in the European population, with individuals with ≥32 repeats having a more than four-fold increased risk (odds ratio 4.25, 95% confidence interval 1.80-12.05). Overall, >92% of expanded alleles harbor CAA interruptions within the CAG tract. Pathogenic expansions at other loci, such as *ATXN3* and *THAP11*, showed more ancestry-specific distributions. Clinically, individuals with pathogenic *ATXN2* and *ATXN3* expansions most often presented with typical PD features but frequently showed earlier disease onset and a strong family history of PD.

This large-scale, multi-ancestry study comprehensively maps the genetic landscape of pathogenic and intermediate repeat expansions in PD. Our findings confirm a length- and structure-dependent risk association for *ATXN2* with PD in the European population and highlight the pleiotropic effects of repeat expansions across the parkinsonian spectrum.

## Introduction

Short tandem repeats are repetitive DNA sequences of 1-6 base pair sequence motifs, comprising at least 6% of the human genome.^1^ Pathogenic repeat expansions underlie a wide spectrum of neurodegenerative and neuromuscular disorders. While repeat expansions in distinct genes may converge on shared phenotypes, individual loci may also give rise to a broad clinical spectrum influenced by repeat length, sequence motif and genomic context.^2^ Uninterrupted full-length pathogenic expansions predominantly cause cerebellar ataxia, whereas intermediate-length alleles or interrupted full-length expansions are more often observed across multiple neurodegenerative diseases, including Parkinson’s disease (PD) and atypical parkinsonism.^3–9^ For example, expansions in Ataxin 2 *(ATXN2)* have been associated with various phenotypes, including spinocerebellar ataxia type 2, motor neuron disease, familial amyloid polyneuropathy and PD, influenced by both repeat length and structural configuration.^10^ Similar pleiotropic effects have been reported for other repeat expansions, such as in Ataxin 3 (*ATXN3)*, Calcium voltage-gated channel subunit alpha 1A (*CACNA1A)* and TATA-Box Binding Protein (*TBP)*, all of which have also been observed in PD.^11–13^ These observations suggest that repeat expansions may contribute more broadly to the pathogenesis of parkinsonism.

Previous studies have been limited to targeted genotyping of a few loci within small or population-specific cohorts, precluding comprehensive assessment of the contribution of repeat expansions to PD at a large, multi-ancestry scale. Consequently, the prevalence, penetrance and phenotypic spectrum of repeat expansions in PD remain incompletely characterized, particularly across ancestrally diverse populations. Furthermore, pathogenic and intermediate repeat length thresholds are inconsistently defined across the literature and may vary with clinical presentation, complicating the interpretation of repeat length–disease relationships. As a result, intermediate alleles, which are known risk factors in other neurodegenerative diseases,^14,15^ may be systematically underrecognized in PD due to variable classification criteria and insufficient statistical power.

Here, we analyzed short-read whole-genome sequencing (WGS) and clinical exome sequencing (CES) data from the Global Parkinson’s Genetics Program (GP2),^16^ the Accelerating Medicines Partnership – Parkinson’s Disease (AMP-PD)^17^ and additional controls from the Alzheimer’s Disease Sequencing Project (ADSP)^18^ to systematically assess established disease-causing repeat expansions detectable by short-read sequencing and evaluate their associations with PD and atypical parkinsonism. We further examined repeat length-disease relationships to determine whether expanded alleles confer PD risk in the European population.

## Materials and Methods

### Ethics declaration

This study was conducted in accordance with the ethical standards of the institutional and national research committees and approved by the ethics committees or institutional review boards at all sites that provided samples and data for this study. Informed consent for study participation was obtained from all participants by each local site. All analyses were conducted using de-identified, anonymized data. No personally identifiable information was accessible to the study investigators.

### Study design and participants

Our study workflow is summarized in **Figure 1**. We integrated data from four primary sources. First, we used WGS data of 18,820 individuals available through GP2 (Release 10, DOI: 10.5281/zenodo.15748014),^19^ with participant recruitment and sample collection described previously.^20,21^ Second, we incorporated WGS data for a total of 6,820 individuals from the Harvard Biomarkers Study (HBS), the Lewy Body Dementia Case-Control Cohort (LBD) and the Parkinson’s Disease Biomarkers Program (PDBP), available through the AMP-PD (Release 4, https://amp-pd.org) platform. Detailed study information is available on the AMP-PD and corresponding study websites. Third, we included 3,221 control samples of European ancestry from ADSP sequenced with PCR-free library preparation and 150 bp paired-end reads (DOI: 10.60859/z6z9-9692).^18^ Lastly, we incorporated clinical exome sequencing (CES) data for 10,454 individuals with PD from the large-scale, international study, PD GENEration^22^ (GP2 Release 8, DOI:10.5281/zenodo.13755496). A summary of cohort demographics for the WGS dataset is provided in **Table 1**.

**Figure 1.**
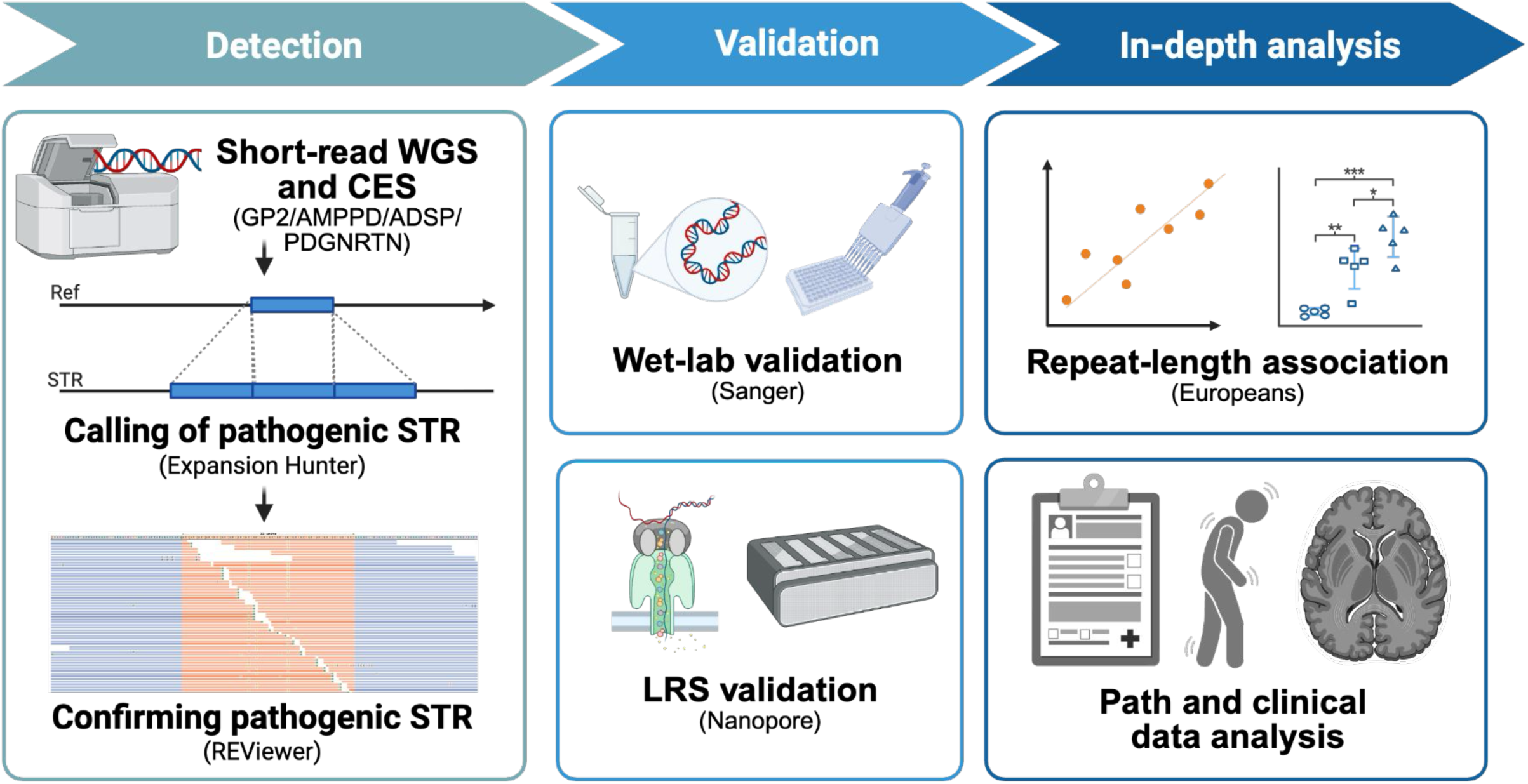
Study Design and Workflow. Figure created with Biorender.

### Whole-genome and clinical exome sequencing datasets

We included a total of 28,861 individuals with whole-genome sequence alignment data available from GP2, AMP-PD (release 4) and ADSP. This cohort included 10,394 healthy controls, 13,738 patients with PD and 4,729 patients with atypical parkinsonism, including progressive supranuclear palsy (PSP), multiple system atrophy (MSA), corticobasal syndrome (CBS) and dementia with Lewy bodies (DLB) (**Table 2**). Potential DLB patients within the LBD cohort were defined as having an intermediate or high likelihood of DLB probability based on McKeith criteria according to the neuropathological data available through AMP-PD. Additionally, we included 10,454 CES samples with clinical exome alignment data as part of GP2 Data Release 8. After removing CES samples identified as genetic duplicates of WGS individuals, a final set of 9,504 CES samples was retained for screening, with the 952 genetic duplicates used to assess the accuracy of CES repeat genotyping.

Both WGS and CES data processing as well as sample and genotype quality control were described previously.^23^ Briefly, we performed single-sample variant calling using DeepVariant v.1.6.1.^24 (^https://github.com/google/deepvariant^)^ followed by joint genotyping of single nucleotide variants and short indels using GLnexus v1.4.3^25^ (https://github.com/dnanexus-rnd/GLnexus). We used KING v.2.3.0^26^ (https://www.kingrelatedness.com, RRID:SCR_009251) to infer relatedness up to the second-degree relatives to confirm known relationships, identify cryptic familial relationships and detect genetic duplicates between WGS and CES datasets. For the WGS dataset, genetic ancestry was determined using GenoTools v1.2.3 (https://github.com/GP2code/GenoTools) with the default settings,^27^ assigning individuals to one of the following genetic ancestries: African-Admixed (AAC), African (AFR), Ashkenazi Jewish (AJ), Latinos and Indigenous People of the Americas (AMR), Complex Admixture (CAH), Central Asian (CAS), East Asian (EAS), European (EUR), Finnish (FIN), Middle Eastern (MDE) and South Asian (SAS).

### Repeat expansion genotyping

We screened disease-causing repeat expansions for which short-read sequencing data can reliably distinguish normal from intermediate and pathogenic alleles.^4,5^ Intermediate and pathogenic repeat size thresholds were obtained from PanelApp (**Supplementary Table 1**). Included repeat expansions met at least one of the following criteria: (i) the intermediate and pathogenic repeat expansion lengths are shorter than 150 bp sequencing read length, allowing accurate discrimination by WGS; or (ii) WGS performance has been validated against the current gold-standard PCR assay with fragment analysis and Sanger sequencing using ExpansionHunter v5.0^28^ (**Supplementary Figure 1**). Although pathogenic expansions in Chromosome 9 Open Reading Frame 72 (*C9orf72)*, Fragile X Messenger Ribonucleoprotein 1 (*FMR1*), Replication Factor C Subunit 1 (*RFC1)* and Fibroblast Growth Factor 14 (*FGF14)* have been reported in PD patients,^29–33^ they did not meet these inclusion criteria and were therefore excluded from our analyses. The resulting genes analyzed were *AR*, *ATN1*, *ATXN1*, *ATXN2*, *ATXN7*, *CACNA1A*, *JPH3* and *TBP* in both WGS and CES datasets. Four additional genes (*ATXN3*, *HTT*, *THAP11*, and *ZFHX3*) were assessed only in the WGS dataset because they were either not captured or yielded low accuracy in the CES data.

We visually inspected repeat length estimates and interruptions for samples carrying intermediate or pathogenic alleles using REViewer.^34^ For the WGS dataset, repeat genotypes supported by ≥2 spanning reads and ≥10 flanking reads that concurred with the spanning read(s) were considered reliable, while for the CES dataset, those supported by ≥5 fully spanning reads were considered reliable. Pathogenic repeats or low-quality genotypes identified in WGS samples were experimentally validated using long-range PCR followed by fragment length analysis or Sanger sequencing of PCR products when DNA was available; otherwise, they were excluded from downstream analyses. Additionally, all predicted intermediate and pathogenic *ATXN3* repeat carriers were validated by Oxford Nanopore sequencing of PCR products for repeat sizing, and those without validation availability were excluded from downstream analyses. The PCR primer sequences and conditions are provided in **Supplementary Table 2**. Concordance between ExpansionHunter genotypes and PCR validation across loci is summarized in **Supplementary Figure 1**. We further evaluated genotype accuracy in the CES dataset using 952 overlapping samples between the two datasets. Overall, all nine loci analyzed in CES showed high concordance in allele genotypes between WGS and CES repeat size (Pearson correlation coefficient 0.97-1; **Supplementary Figure 2**).

### Statistical analysis

#### Carrier frequency estimate (1 in x)

We computed the pathogenic and intermediate carrier frequencies stratified by phenotypes and ancestries. The carrier frequency was expressed as a 1 in x ratio, where x represents the total number of unrelated genomes divided by the number of pathogenic or intermediate alleles. Confidence intervals (CI) for the carrier frequencies were computed using the Wilson score method:

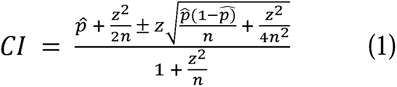

where *p* = the total number of pathogenic or intermediate alleles divided by the total number of unrelated genomes (*n*); *z* = 1.96.

#### Repeat-length association with PD

To evaluate PD risk across a range of repeat lengths at each locus and identify the minimum thresholds associated with increased disease risk, we performed threshold-based association analyses using Firth’s penalized logistic regression on the five loci (*ATXN1*, *ATXN2*, *CACNA1A*, *HTT*, and *TBP*) where intermediate alleles were observed. Cumulative repeat-length bins were defined by their lower limits, with allele status within each bin coded as a binary variable (1 for expanded; 0 for non-expanded). For pairs of related individuals, we retained only one individual per pair, preferentially keeping PD patients over controls for the downstream analysis. The model adjusted for sex and the first five principal components, with maximum iterations set to 1000 and other parameters set to default. To account for multiple testing, we applied a study-wide Bonferroni correction across the 44 repeat-length bins evaluated across the five loci, resulting in a significance threshold of *P* < 0.0011. The analyses were performed using the logistf package (v1.23) in R (v4.4.2) using the European WGS dataset. Analyses were not performed in other ancestries due to limited sample sizes or the lack of controls.

## Results

### Pathogenic repeat expansions in PD and atypical parkinsonism

A total of 28,861 individuals with short-read WGS data from GP2, AMP-PD and ADSP were included in the analysis, comprising 13,738 patients with PD, 4,729 patients with atypical parkinsonism (DLB, CBS, MSA and PSP) and 10,394 healthy controls across 11 genetic ancestries. Both pathogenic and intermediate repeat expansions were detected across multiple loci and ancestries (**Figure 2**; **Figure 3; Supplementary Table 3**).

**Figure 2.**
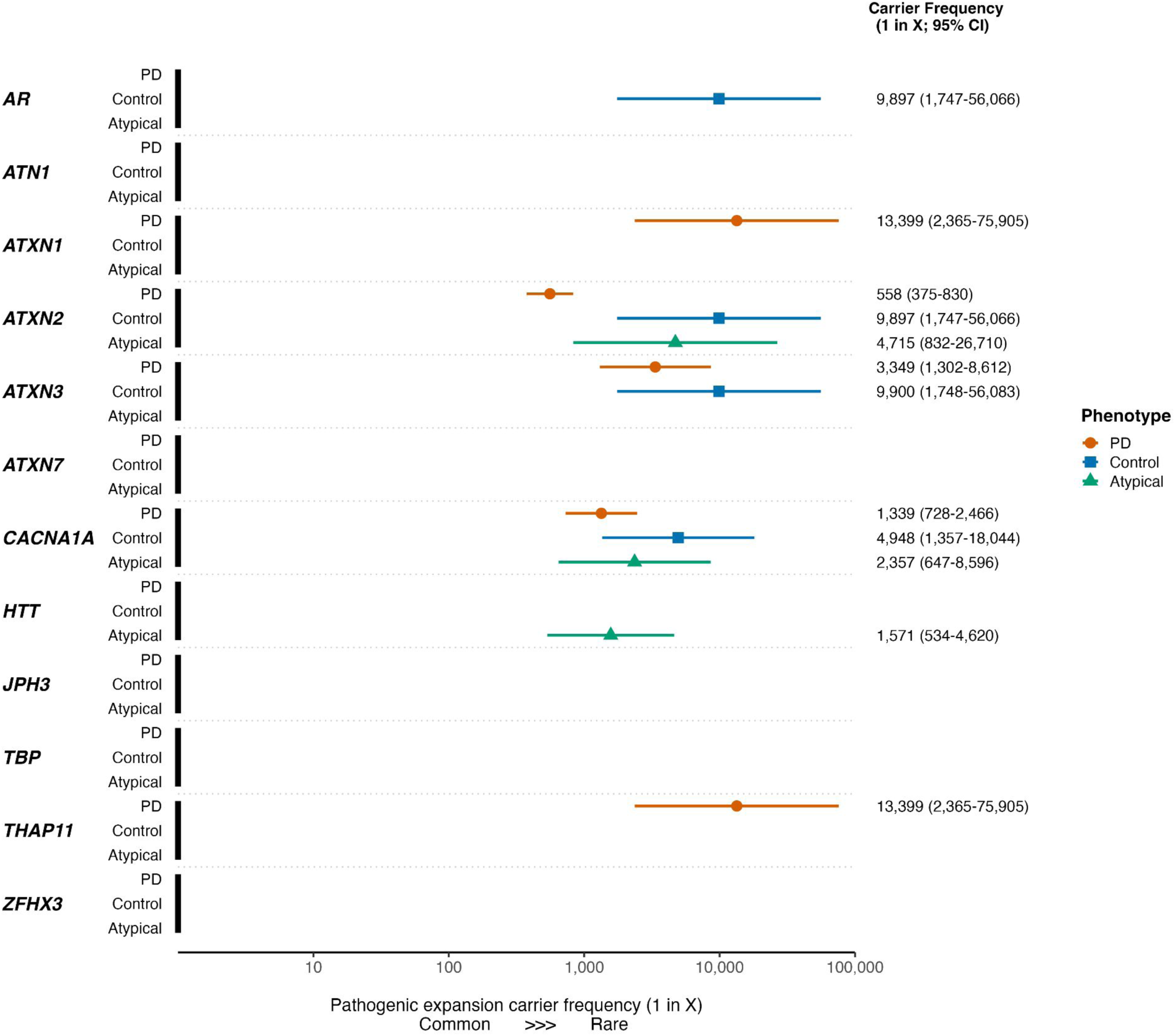
Forest plot of pathogenic expansion carrier frequencies across phenotypes in the WGS dataset. Pathogenic expansion carrier frequency (expressed as 1 in x individuals; 95% confidence interval) is shown across phenotypes for 12 loci. Points represent the estimated carrier frequency (1 in x) and horizontal lines indicate 95% confidence intervals; an empty line indicates that no carriers were detected. Symbols indicate phenotype (PD = Parkinson’s Disease, Atypical = atypical parkinsonism). See Supplementary Table 3 for the exact number of individuals included (N may vary slightly across loci due to data quality control and filtering).

**Figure 3.**
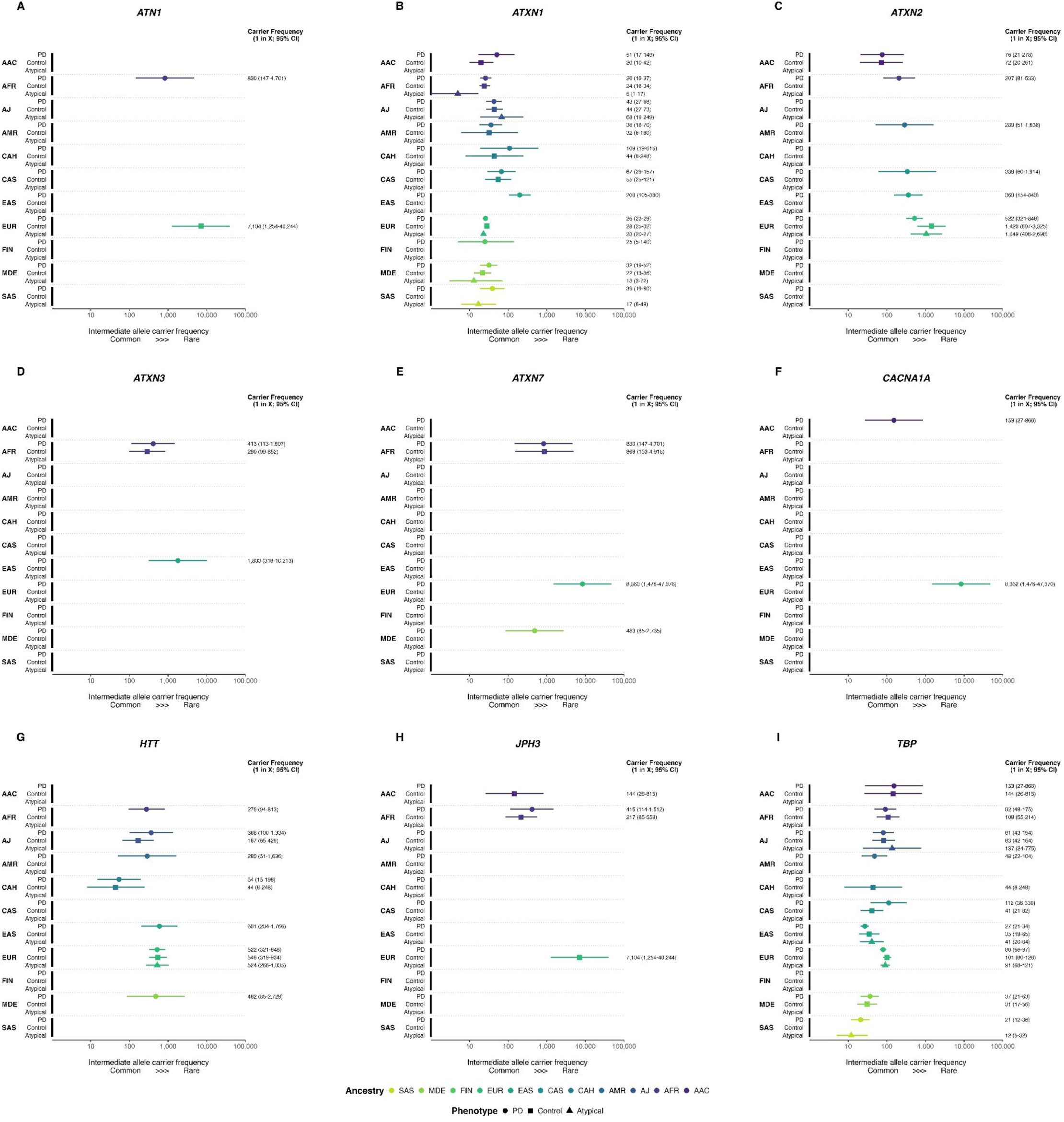
Forest plot of intermediate allele carrier frequencies across phenotypes and genetic ancestries in the WGS dataset. Intermediate repeat allele carrier frequencies (expressed as 1 in x individuals; 95% confidence interval [CI]) are shown across ancestries and phenotypes for nine loci: ATN1 (A), ATXN1 (B), ATXN2 (C), ATXN3 (D), ATXN7 (E), CACNA1A (F), HTT (G), JPH3 (H), and TBP (I). Points represent estimated carrier frequencies, and horizontal lines indicate 95% CIs; an empty line indicates that no carriers were detected. Colors denote genetic ancestry: African-Admixed (AAC), African (AFR), Ashkenazi Jewish (AJ), Latinos and Indigenous People of the Americas (AMR), Complex Admixture (CAH), Central Asian (CAS), East Asian (EAS), European (EUR), Finnish (FIN), Middle Eastern (MDE), and South Asian (SAS). Symbols indicate phenotype (PD = Parkinson’s Disease, Atypical = atypical parkinsonism). See Table 1 for the total number of individuals included; baseline numbers (N) may vary slightly across loci due to data quality control and filtering.

Pathogenic repeat expansions were identified in 67 individuals, including 56 with PD, 1 with DLB, 2 with MSA, 1 with PSP, 2 with CBS and 5 healthy controls at seven loci (**Table 2; Supplementary Table 3**). *ATXN2* expansions were most frequently observed, with carriers detected across four ancestries: AFR (1/833 PD, 0.12%; 1/886 controls, 0.11%), EAS (12/1859 PD, 0.65%), EUR (13/8589 PD, 0.15%; 1/2669 DLB, 0.04%) and MDE (1/492 PD, 0.20%).

Similarly, pathogenic *CACNA1A* expansions were also detected across multiple ancestries, including AMR (1/294 PD, 0.34%), EAS (5/1857 PD, 0.27%; 1/185 MSA, 0.54%; 1/390 controls, 0.26%) and EUR (4/8590 PD, 0.05%; 1/983 PSP, 0.10%; 1/7403 controls, 0.01%). Pathogenic *ATXN3* expansions were observed only in AFR (3/830 PD, 0.36%; 1/888 controls, 0.11%) and EAS (1/1859 PD, 0.05%). Additional loci with pathogenic expansions included *AR* (EUR, 1/7403 controls, 0.01%), *ATXN1* (EUR, 1/8591 PD, 0.01%), *HTT* (EUR, 2/166 CBS, 1.20%; 1/387 MSA, 0.26%) and *THAP11* (EAS, 1/1857 PD, 0.05%). Notably, the expanded *ATXN1* allele exhibited CAA interruptions and was therefore not considered pathogenic for ataxia. No pathogenic repeat expansions were detected in *ATN1*, *ATXN7*, *JPH3*, *TBP* and *ZFHX3*. Among the 9,504 non-overlapping PD patients with CES data (no genetic ancestry prediction available), pathogenic repeat expansions were identified in *ATXN2* (*n*=6, 0.06%), *CACNA1A* (*n*=1, 0.01%) and *AR* (*n*=6, 0.06%).

Among the carriers of pathogenic repeat expansions, *ATXN2* expansions co-segregated with PD in three families, including one of EAS ancestry and two of EUR ancestry (**Supplementary Figures 3A-C**). Another individual with a *ATXN2* repeat expansion reported an affected sibling with PD, who underwent genetic testing outside this study and was confirmed to carry an *ATXN2* repeat expansion as well (**Supplementary Figures 3D**). In addition to these families exhibiting a pure PD phenotype, we also identified a family of EAS ancestry showing phenotypic heterogeneity driven by intergenerational instability of the *ATXN2* CAG tract. Specifically, the father was diagnosed with PD and carried an uninterrupted expansion of 38 repeats, while his son was diagnosed with cerebellar ataxia and carried a larger uninterrupted expansion of 42 repeats (**Supplementary Figure 3E**). In addition to *ATXN2*, we also identified a family of EAS ancestry showing pathogenic *ATXN3* expansion segregating with the disease (**Supplementary Figure 3F**).

### Intermediate repeat frequency in different populations

In contrast to rare pathogenic repeat expansions, intermediate alleles were substantially more prevalent and broadly more distributed across loci and ancestries (**Figure 3**). We observed *ATXN1*, *HTT* and *TBP* intermediate alleles exhibiting broad cross-ancestry distributions with comparable carrier frequencies between PD patients and controls, suggesting limited phenotype-specific enrichment. *ATXN1* intermediate alleles showed higher frequencies in EUR (1 in 26 for PD; 1 in 28 for control), SAS (1 in 39 for PD) and MDE (1 in 32 for PD; 1 in 22 for control) ancestries, compared with EAS (1 in 200 for PD). By contrast, *TBP* intermediate alleles seem to be more frequent in EAS (1 in 27 for PD; 1 in 35 for control), MDE (1 in 37 for PD; 1 in 31 for control) and SAS (1 in 21 for PD) ancestries and less common in AAC (1 in 153 for PD; 1 in 144 for control), AFR (1 in 92 for PD; 1 in 108 for control) and AJ (1 in 81 for PD and 1 in 108 for control) ancestries. Intermediate alleles in *ATXN2* were detected across six ancestries (AAC, AFR, AMR, CAS, EAS and EUR) and occurred predominantly in PD patients. Notably, in the European population, *ATXN2* intermediate alleles were more frequent in PD patients (1 in 522) than in controls (1 in 1,420).

Comparatively, we found that intermediate alleles in *ATN1*, *ATXN3*, *ATXN7*, *JPH3* and *CACNA1A* were rarer and highly ancestry-specific. Specifically, *ATXN3* intermediate alleles were primarily observed in AFR (1 in 413 for PD; 1 in 209 for controls) and EAS (1 in 1,803 for PD) ancestries. Similarly, intermediate *ATXN7* alleles were largely restricted to AFR (1 in 830 for PD; 1 in 868 for controls) and MDE (1 in 483 for PD) ancestries. Intermediate *CACNA1A* alleles were detected only in PD patients of EUR (1 in 8362) and AAC (1 in 153) ancestry, whereas *JPH3* intermediate alleles were primarily limited to the AFR (1 in 415 for PD; 1 in 217 for control) and AAC (1 in 144 for control) ancestries. Finally, *ATN1* intermediate alleles were rare and detected almost exclusively in the EUR (1 in 7104 for control) and AFR (1 in 830 for PD) ancestries.

### Repeat-length disease association analyses

We next evaluated the association between repeat expansions and PD risk. We performed threshold-based association analyses across five genes (*ATXN1, ATXN2, CACNA1A, HTT* and *TBP*) where intermediate alleles were observed, with the aim of identifying the minimum repeat-length thresholds associated with increased disease risk. Using Firth’s penalized logistic regression adjusted for sex and the first five principal components in 7,963 European PD patients and 6,757 controls, we observed a significant association between expanded *ATXN2* repeat length and PD risk across multiple length thresholds after Bonferroni correction for multiple testing (**Figure 4**; **Supplementary Table 4**). At a threshold of ≥32 repeats (26 patients, 5 controls), carriers had a more than four-fold increased risk of PD compared with non-carriers (odds ratio [OR] = 4.25, 95% CI: 1.80 to 12.05, *P* = 0.00057). Stronger effect estimates were observed at higher cutoffs, including ≥33 repeats (17 patients, 1 control; OR = 10.95, 95% CI: 2.71 to 100.07, *P* = 0.00017) and ≥34 repeats (14 patients, 0 controls; OR = 27.55, 95% CI: 3.59 to 3540.26, *P* = 0.00012) although the risk estimate was statistically unstable due to zero observations in the controls. There was no association between repeat length and age at onset (AAO) (**Supplementary Figure 5**). In contrast to *ATXN2*, no significant associations with PD risk were detected for repeat expansions in *HTT*, *ATXN1*, *CACNA1A*, or *TBP* at any threshold examined in the European dataset (**Supplementary Table 4**). Of note, statistical power was limited for *CACNA1A* due to its low carrier frequency in this study. Taken together, these findings support a repeat length-dependent effect of *ATXN2* expansions on PD risk.

**Figure 4.**
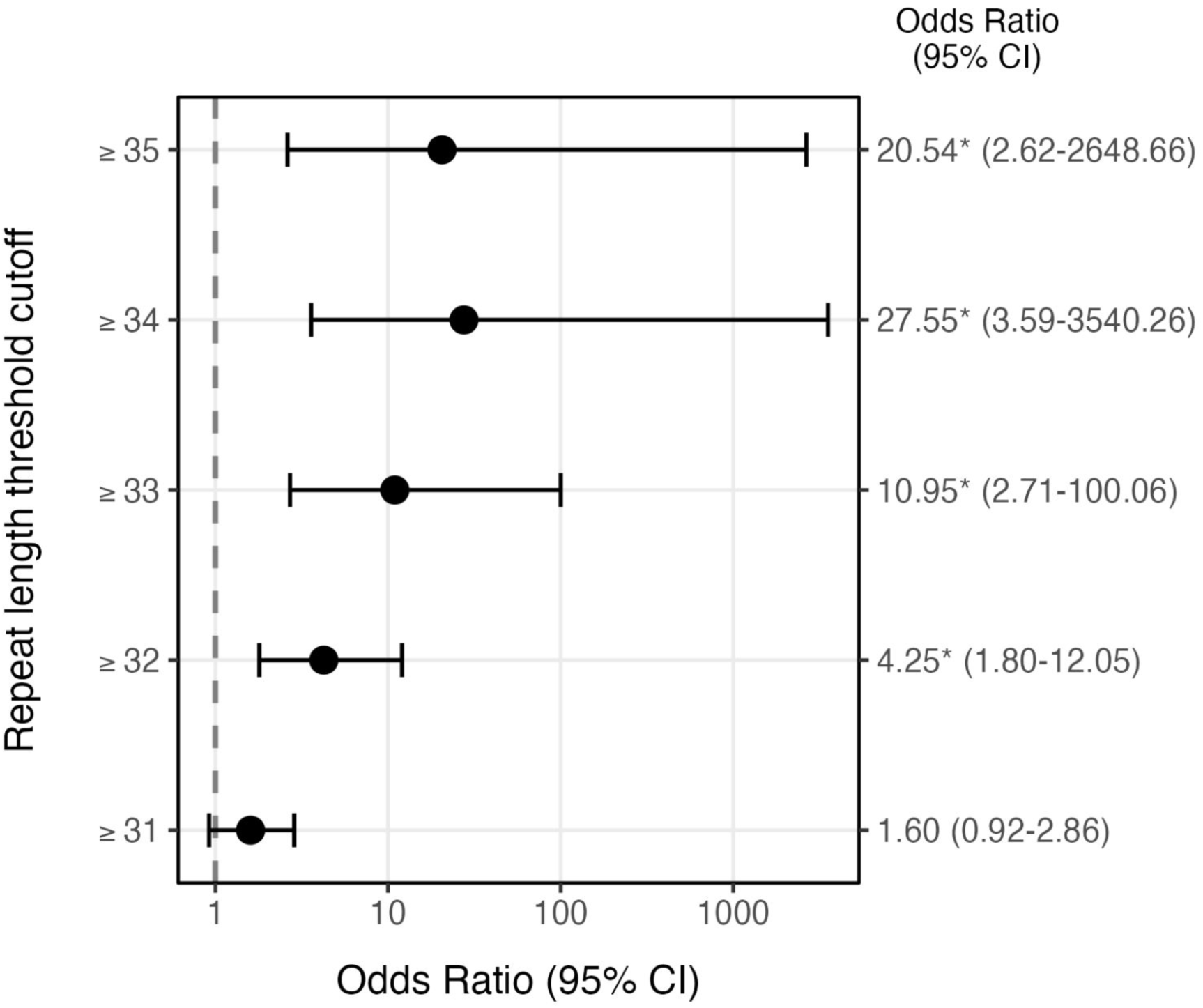
Threshold-based association analysis of ATXN2 repeat length and Parkinson’s disease risk in the European population. Forest plot showing odds ratios and 95% confidence intervals (CIs) for Parkinson’s disease across cumulative ATXN2 repeat-length bins (≥31 to ≥35 repeats). Points represent odds ratio estimates and horizontal lines indicate 95% CIs; the dashed vertical line denotes odds ratio of 1; * indicate significant P value after Bonferroni correction (P < 0.0011).

### Repeat motif structure in expanded alleles

In addition to repeat length, we also examined repeat structure by inspecting sequence alignments or Sanger sequencing of pathogenic repeat expansions detected in this study. Given the observed *ATXN2* length-based association in the European population, we additionally characterized the repeat motif in individuals harboring 32–34 *ATXN2* repeats (**Table 2**; **Supplementary Table 5**). Among European carriers of expanded alleles (≥32 repeats) across phenotypes, 92.7% (38/41) contained one or more CAA interruptions within the CAG tract. The frequency of interruptions differed across ancestries, occurring in 76.5% (13/17) of EAS carriers and 66.7% (4/6) of AFR carriers. All expanded allele carriers of AAC (4/4), AMR (1/1), CAS (1/1) and MDE (1/1) ancestry also harbored at least one CAA interruption; however, these estimates should be interpreted cautiously given the limited sample sizes in non-European ancestries. Similarly, 88.5% (23/26) of *ATXN2* expanded allele carriers identified from non-overlapping PD patients with CES data contained at least one CAA interruption.

Across other loci, repeat motif analysis revealed locus-specific interruption patterns. The pathogenic *THAP11* allele in a PD patient of EAS ancestry showed seven CAA interruptions distributed throughout the expanded CAG tract, with the 3’ uninterrupted CAG tract of 10 repeats (**Supplementary Figure 4**). The single pathogenic *ATXN1* carrier harbored four CAA interruptions. All five pathogenic *ATXN3* alleles exhibited the canonical CAA interruption pattern, whereas all 14 pathogenic *CACNA1A* alleles consisted exclusively of pure CAG repeats. Finally, three pathogenic *HTT* alleles exhibited the typical canonical (CAG)_n_-CAA-CAG-CCA-(CCG)_n_ configuration.

### Clinical characteristics of confirmed pathogenic repeat expansion carriers

Individual-level clinical and demographic characteristics of all pathogenic repeat expansion carriers are summarized in **Table 2**.

Among 35 individuals with *ATXN2* repeat expansions, 33 had PD without any atypical features, one patient was diagnosed with DLB and one was an unaffected carrier. AAO ranged from 38 to 67 years (median 50 years; missing for *n*=5), with 53.6% (15/28) having an early disease onset (≤50 years). A positive family history of PD was reported by 87.9% of individuals (29/33; missing for *n*=1), including several families with multiple affected individuals (**Supplementary Figure 3**). Neuropathological data were available for one individual with PD demonstrating Lewy body pathology. In addition, one individual had DLB, and one unaffected individual in their 70’s had no reported family history of PD. Characteristics of individuals with intermediate *ATXN2* repeat expansions are summarized in Supplementary Table 5.

Among five *ATXN3* repeat expansion carriers, four individuals had early onset PD (AAOs 35–42 years), three of whom (75.0%) reported a positive family history of PD. The fifth carrier was an unaffected control (age 40-44 years) without a reported family history of PD. Pathogenic *ATXN1* expansions were rare and only identified in a single individual with PD (AAO 55-59 years) without a reported family history of PD.

For *CACNA1A* repeat expansions, most carriers (11/15) had classical PD without atypical features, with AAO ranging from 31 to 65 years (median 47 years; missing for *n*=2), and 70.0% (7/10) reported a positive family history of PD (missing for *n*=1). Neuropathological data were available for one individual, demonstrating brain stem Lewy body pathology, and α-synuclein seeding activity in cerebrospinal fluid was positive in another individual with PD. Two individuals had atypical parkinsonism, one with PSP (AAO 65-69 years) and the other with MSA-P (AAO 60-64 years). Neither of them reported a family history of PD. Two additional individuals were unaffected, one male in his late 80’s and one female of unknown age.

Pathogenic *HTT* expansions were identified only in individuals with atypical parkinsonian phenotypes. These included one female with neuropathologically confirmed MSA (AAO: 60-64 years) and two males with CBS (AAOs 65-69 and 70-74 years), one of whom also had pathological confirmation of corticobasal degeneration. None of the *HTT* pathogenic repeat carriers reported a family history of PD. Furthermore, a single female carrier of pathogenic *THAP11* repeat had early-onset PD (AAO 40-44 years) without a reported family history.

Finally, seven male carriers of pathogenic *AR* expansions were identified, consistent with an X-linked mode of inheritance. Six had PD, with AAO ranging from 59 to 72 years (median 63 years), and none reported a family history of PD (missing for *n*=1). One additional carrier was an unaffected individual in their 50’s who reported a positive family history of PD.

## Discussion

While pathogenic expansions in *ATXN2*, *ATXN3*, *CACNA1A*, *TBP* and *HTT* are classically associated with distinct neurodegenerative syndromes, they have also been observed in PD and atypical parkinsonism, predominantly *ATXN2* in EAS and EUR and *ATXN3* in AFR populations.^4,6,12,29–31,35–40^ Our multi-ancestry study corroborates and extends these observations by demonstrating population-specific differences in repeat expansion frequencies and strengthening evidence for the pleiotropic effects of repeat expansions across the neurodegenerative disease spectrum.

Consistent with prior reports,^11,41^ *ATXN2* expansions were the most frequently detected in PD among the genes evaluated. Furthermore, their presence across four ancestries (AFR, EAS, EUR and MDE) indicates a broader cross-ancestry contribution to PD compared to the other genes analyzed. Notably, we observed a higher frequency of *ATXN2* pathogenic expansions in EAS ancestry compared with other ancestries and reported the first identification of pathogenic expansions in PD patients of MDE and AFR ancestry. By contrast, pathogenic expansions in *ATXN3* were confined to AFR and EAS ancestries mostly in PD, while a *THAP11* expansion was found only in a PD patient of EAS ancestry. Pathogenic *THAP11* expansions have previously been reported in patients with ataxia and intellectual disability.^42,43^ By identifying *THAP11* pathogenic expansion in PD, we highlight a potential extension of the phenotypic spectrum associated with *THAP11* repeat expansions. Additionally, pathogenic *CACNA1A* expansions were detected in both PD and atypical parkinsonism, whereas *HTT* expansions were observed only in atypical parkinsonism. Collectively, these findings demonstrate both cross-ancestry occurrences and ancestry-specific enrichments of established pathogenic repeat expansions across the parkinsonian phenotypic spectrum.

Intermediate alleles were substantially more prevalent than pathogenic expansions and were broadly distributed across loci and ancestries, consistent with observations in the general populations.^44–46^ Our estimated intermediate allele carrier frequencies across ancestries show similar patterns to those previously reported,^44^ where *ATXN1* intermediate alleles were more frequently observed in the EUR and SAS ancestries, *HTT* in EUR and AFR ancestries, and *TBP* in EAS ancestry. Conversely, intermediate alleles in *ATN1*, *ATXN3*, *ATXN7*, *CACNA1A* and *JPH3* were comparatively rare and showed ancestry-specific distributions. For instance, pathogenic and intermediate *ATXN3* expansions were detected exclusively in AFR and EAS ancestries, and were notably absent in EUR ancestry consistent with previous studies.^41,44,47^ This restricted distribution likely reflects a limited number of founder haplotypes.^48^ Moreover, *ATXN3* expansions in PD have been described only in patients of AFR and EAS ancestry.^13,47,49–51^ Collectively, these observations suggest that genetic background may influence the phenotypic expression of repeat expansions and contribute to ancestry-specific disease manifestations. This warrants further investigation into population-specific cis- and trans-acting genetic modifiers of repeat expansions, which might underlie differences in prevalence across populations.

The pathogenicity of repeat expansions is often modulated by repeat length, structure and genomic context.^2^ Our results provide evidence for a repeat-length-dependent association between *ATXN2* expansions and PD risk, extending to intermediate length repeat expansions, comparable to what was previously shown for amyotrophic lateral sclerosis.^15^ Increasing *ATXN2* repeat size was associated with progressively higher risk across multiple thresholds in the EUR population, with carriers of ≥32 repeats showing significantly increased risk of PD (OR = 4.25) and larger effect estimates observed at higher repeat lengths. Although estimates at the largest thresholds were imprecise due to low carrier counts, the overall pattern supports a graded relationship between *ATXN2* repeat length and PD risk. Notably, comparable length-dependent associations were not observed for expansions at *ATXN1*, *HTT*, or *TBP*, suggesting gene-specific mechanisms rather than a generalized effect of CAG expansion burden. Furthermore, we found no PD enrichment among intermediate alleles in *ATXN1*, *HTT* and *TBP* that were previously suggested as having reduced penetrance in ataxia and Huntington’s disease.^12,52,53^ Consequently, the role of these alleles in PD appears limited, and their clinical significance should be interpreted cautiously.

In addition to repeat length, interruption motifs within CAG tracts, such as CAA or CAT, are relatively common and known to influence the phenomenon of anticipation through stabilizing the repeat transmission, thereby influencing disease risk and phenotype expression in polyglutamine diseases.^10^ In particular, interrupted *ATXN2* repeats have been associated with amyotrophic lateral sclerosis and parkinsonism rather than the classical ataxia phenotype.^6,8,36^ Consistent with prior reports, among EUR carriers with ≥32 repeats, more than 90% harbored at least one interruption, with similarly high albeit more variable frequencies across other ancestries, supporting the hypothesis that the repeat structure contributes to the parkinsonian presentation associated with *ATXN2* expansions.

Repeat structure may also modulate the pathogenicity of repeat expansions at other loci. Previous studies have suggested that *THAP11* repeat expansion pathogenicity is primarily driven by the length of the uninterrupted 3’ CAG tract, with ataxia patients harboring ≥32 pure CAG repeats at this position, whereas unaffected individuals typically carry 4–16 uninterrupted repeats.^42,43^ Consistent with this, the pathogenic *THAP11* allele identified in an East Asian PD patient exhibited multiple CAA interruptions and an uninterrupted 3′ tract of 10 CAG units, supporting the conclusion that repeat configuration influences phenotypic expression. Conversely, pathogenic *ATXN3* expansions consistently exhibited the canonical CAA interruption pattern also seen in ataxia patients,^54^ while *CACNA1A* alleles comprised strictly pure CAG repeats, suggesting that the effect of repeat structure on phenotype might be locus-specific.

Among the loci analysed, *ATXN2* and *ATXN3* have been most consistently linked to PD phenotypes.^6,7,35–37,39^ Notably, carriers of pathogenic expansions in both genes within our cohort showed a strikingly high proportion of early AAO. Furthermore, *ATXN2* carriers frequently reported a positive family history of PD, reinforcing the relevance of *ATXN2* expansions in familial and early-onset PD. Similarly, *CACNA1A* expansions, which have not been commonly associated with classical PD phenotypes,^55^ were predominantly observed in individuals with early-onset disease and often with a positive family history of PD. These observations broaden the phenotypic spectrum of repeat expansions and underscore the complexity of their clinical interpretation in PD. In parallel, our broad genomic screening approach also increases the possibility of incidental findings. Identifying highly penetrant pathogenic expansions associated with other severe neurodegenerative disorders in individuals presenting with parkinsonism, or even in asymptomatic controls, raises important ethical considerations regarding informed consent, genetic counseling and the return of unexpected results. This complexity is further compounded when considering risk-associated intermediate length repeat expansions. Clinically, such findings emphasize the need for careful (reverse) phenotyping to distinguish true pleiotropy from misdiagnosis and to ensure accurate diagnostic and prognostic counseling.

While our study represents one of the largest multi-ancestry assessments of repeat expansions in PD, it has several limitations. First, some populations were represented by relatively small sample sizes. In addition, molecular validation was feasible only for the subset of individuals with available DNA samples, potentially resulting in underascertainment of expansion carriers. Larger and more diverse WGS datasets will therefore be crucial to replicate these findings and to refine estimates of expansion frequencies and associated risk beyond EUR ancestry.^56,57^ Second, short-read WGS cannot reliably size expansions exceeding the read length, highlighting the need for future large-scale long-read sequencing to resolve full repeat length and structures for very large expansions and to improve their characterization in PD. Third, clinical data were largely restricted to primary clinical diagnoses, and detailed information on family history beyond PD was also limited. Deeper clinical and biological phenotyping, including specific neurological features, neuroimaging and biomarkers, are essential for robust genotype-phenotype correlations.

Taken together, our findings provide a comprehensive landscape of 12 repeat expansions in PD and atypical parkinsonism, highlighting both full-length pathogenic alleles and more frequent intermediate alleles that may modify disease risk across diverse populations. The observed differences in carrier frequency across ancestries emphasize the importance of including diverse populations in genetic studies to fully capture the spectrum of repeat-mediated risk. In addition, our results emphasize the importance of considering certain repeat expansions, i.e., *ATXN2, ATXN3* and *CACNA1A* in the genetic evaluation of PD and, to an extent, atypical parkinsonian syndromes, while acknowledging the differences in prevalence across ancestries.

## Data availability

Data used in the preparation of this article were obtained from Global Parkinson’s Genetics Program (GP2; https://gp2.org). Specifically, we used Tier 2 data from GP2 Release 8 (https://doi.org/10.5281/zenodo.13755496) and GP2 Release 10 (https://doi.org/10.5281/zenodo.15748014). Additional whole-genome sequencing data used in the preparation of this article were obtained from the Accelerating Medicine Partnership® (AMP®) Parkinson’s Disease (AMP PD) Knowledge Platform. For up-to-date information on the study, visit https://www.amp-pd.org. Qualified researchers are encouraged to apply for direct access to the data through AMP-PD. The GP2 and AMP-PD datasets analysed during the current study are available through AMP-PD (https://amp-pd.org). Data from Alzheimer’s Disease Sequencing Project (NG00067.v10, August 15, 2023, DOI: 10.60859/z6z9-9692) used in this study were prepared, archived, and distributed by the National Institute on Aging Alzheimer’s Disease Data Storage Site (NIAGADS) at the University of Pennsylvania (U24-AG041689), funded by the National Institute on Aging. The full acknowledgement statement for the ADSP can be found at: https://dss.niagads.org/datasets/ng00067/.

All code generated for this article and the identifiers for all software programs and packages used, are available on GitHub (https://github.com/GP2code/GP2-STR) and were given a persistent identifier via Zenodo (10.5281/zenodo.20432080).

## Acknowledgments

We would like to thank Max Brand, Frauke Hinrichs, Heike Pawlack and Alicia Wenghöfer for technical assistance in experimental validation.

This project was supported by the Global Parkinson’s Genetics Program (GP2; https://gp2.org). GP2 is funded by the Aligning Science Against Parkinson’s (ASAP) (https://ror.org/03zj4c476) Initiative and implemented by The Michael J. Fox Foundation for Parkinson’s Research (https://ror.org/03arq3225). For a complete list of GP2 members see doi.org/10.5281/zenodo.7904831.

The AMP® PD program is a public-private partnership managed by the Foundation for the National Institutes of Health and funded by the National Institute of Neurological Disorders and Stroke (NINDS) in partnership with the Aligning Science Across Parkinson’s (ASAP) initiative; Celgene Corporation, a subsidiary of Bristol-Myers Squibb Company; GlaxoSmithKline plc (GSK); The Michael J. Fox Foundation for Parkinson’s Research; Pfizer Inc.; AbbVie Inc.; Sanofi US Services Inc.; and Verily Life Sciences. ACCELERATING MEDICINES PARTNERSHIP and AMP are registered service marks of the U.S. Department of Health and Human Services. Clinical data used in the preparation of this article were obtained from the MJFF-sponsored LRRK2 Cohort Consortium (LCC). For up-to-date information on the study, visit www.michaeljfox.org./lcc. The LRRK2 Cohort Consortium is coordinated and funded by The Michael J. Fox Foundation for Parkinson’s Research. The investigators within the LCC provided data, but did not participate in the analysis or writing of this report. The full list of LCC investigators can be found at www.michaeljfox.org/lccinvestigators. Clinical exome sequencing data was contributed by PD GENEration, a flagship initiative of the Parkinson’s Foundation in partnership with Aligning Science Across Parkinson’s (ASAP) and the Global Parkinson’s Genetics Program (GP2). Additional financial and in-kind support comes from the Parkinson’s community – industry partners, nonprofit organizations, and individuals whose lives have been touched by Parkinson’s. For up-to-date information on the study, visit https://www.parkinson.org/pdgeneration.

This research was supported in part by the Intramural Research Program of the National Institutes of Health (NIH). The contributions of the NIH author(s) are considered Works of the United States Government. The findings and conclusions presented in this paper are those of the author(s) and do not necessarily reflect the views of the NIH or the U.S. Department of Health and Human Services.

Biospecimens used in the analyses presented in this article were obtained from the Northwestern University Movement Disorders Center (MDC) Biorepository. As such, the investigators within MDC Biorepository contributed to the design and implementation of the MDC Biorepository and/or provided data and collected biospecimens but may not have participated in the analysis or writing of this report. MDC Biorepository investigators include Rizwan Akhtar MD, PhD; Tanya Simuni MD; Dimitri Krainc, MD, PhD; Puneet Opal MD, PhD; Joanna Blackburn MD; Monika Szela MHA; and Lisa Kinsley MS, CGC. A gift from the Malkin family generously supported the work of the MDC Biorepository.

## Appendix 1

GP2 Banner authorship

