## Supplementary Figures for "Repeat expansions in Parkinson’s disease and parkinsonism across ancestries: insights from a global genetic cohort"

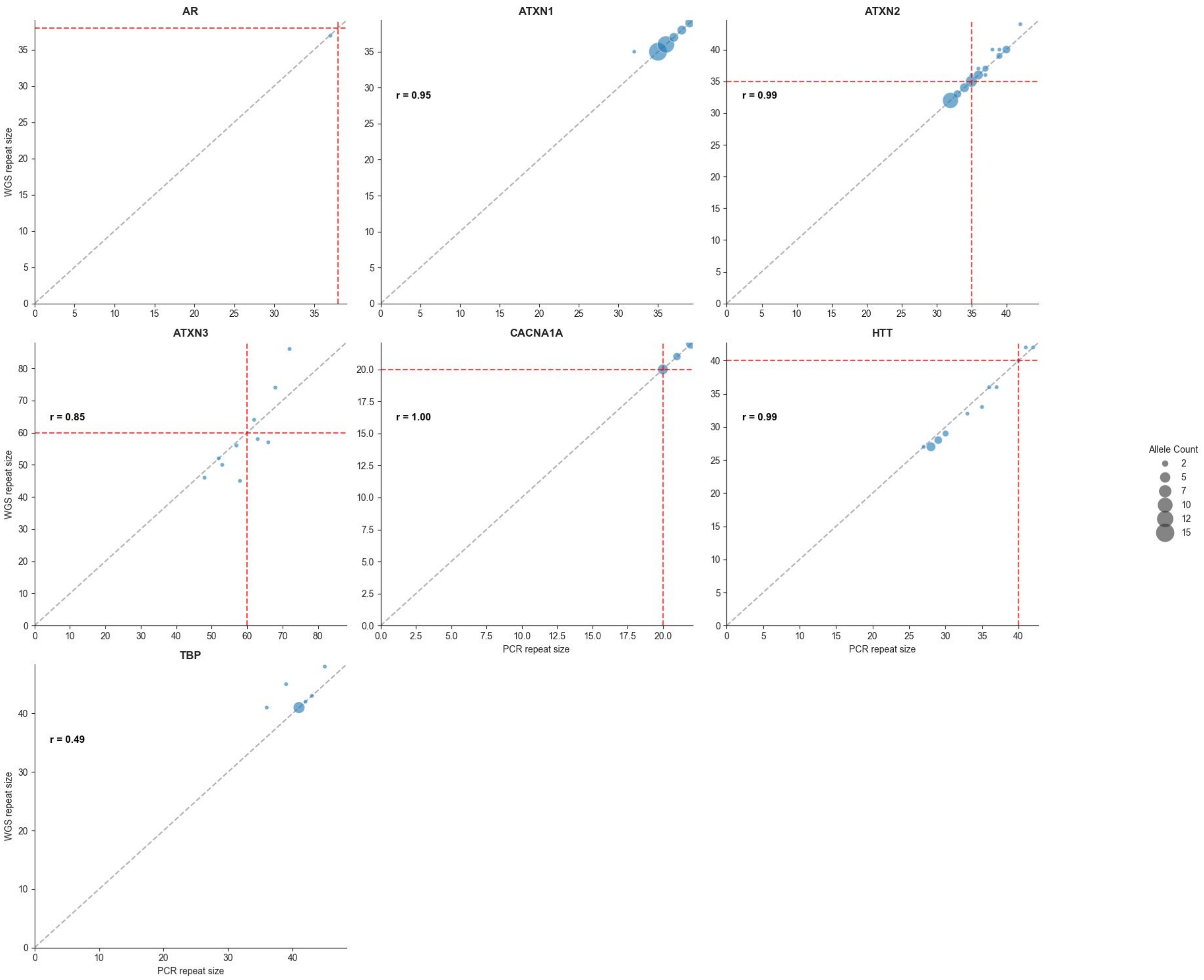


**Supplementary Figure 1. Repeat size correlation by locus between PCR repeat size and repeat expansion detection using whole-genome sequencing (WGS).** Scatter plots show PCR repeat sizes on the x axes and WGS ExpansionHunter repeats sizes on y axes, and , and r represent the Pearson correlation coefficient. Red dashed lines represent the pathogenic thresholds. The size of the dot represents the total number of alleles tested.

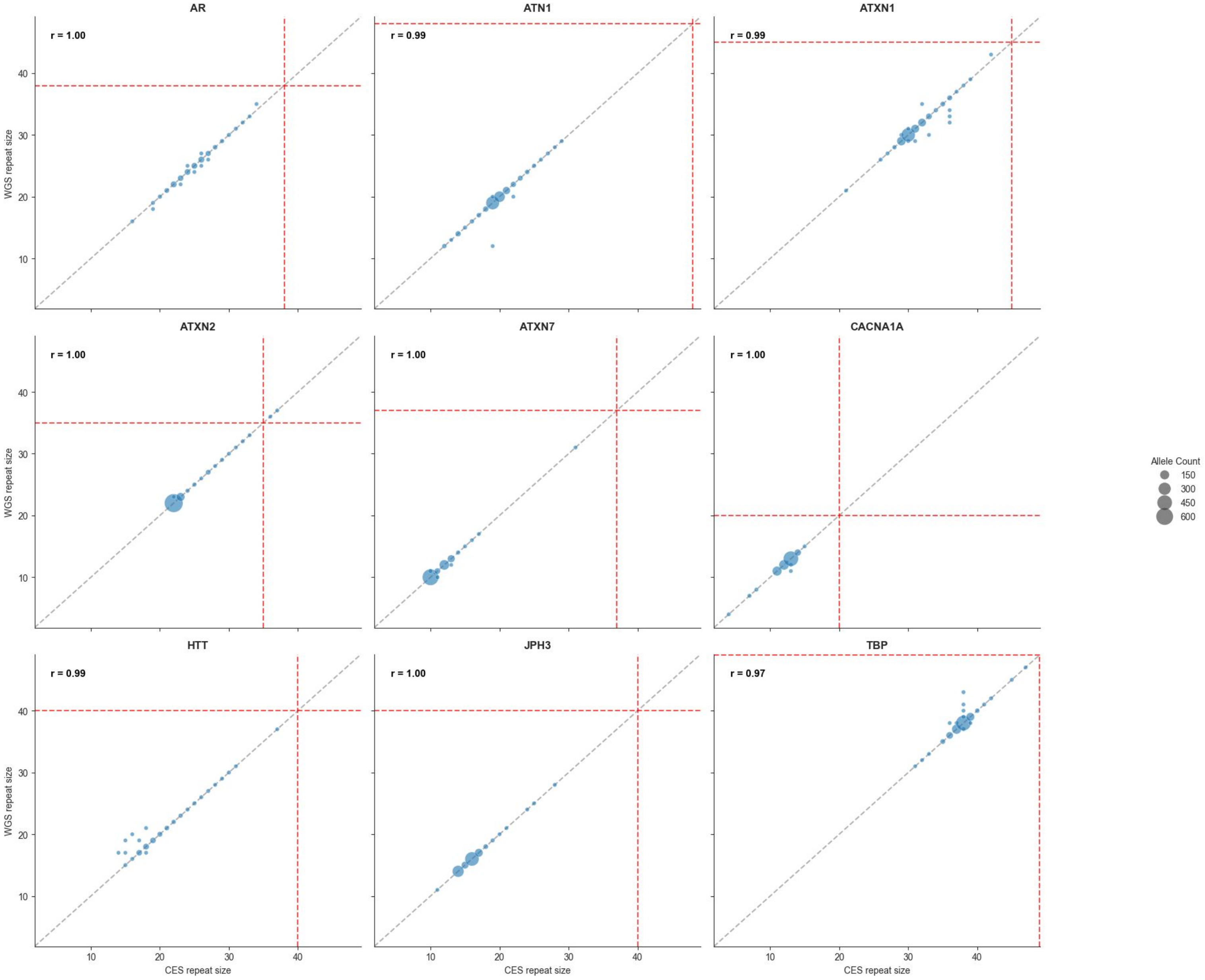


**Supplementary Figure 2. Repeat size correlation by locus between repeat expansion detection using whole-genome (WGS) and clinical exome sequencing (CES).** Scatter plots show CES ExpansionHunter repeats sizes on the x axes and WGS ExpansionHunter repeats sizes on y axes, and r represent the Pearson correlation coefficient. Red dashed lines represent the pathogenic thresholds. The size of the dot represents the total number of alleles tested.

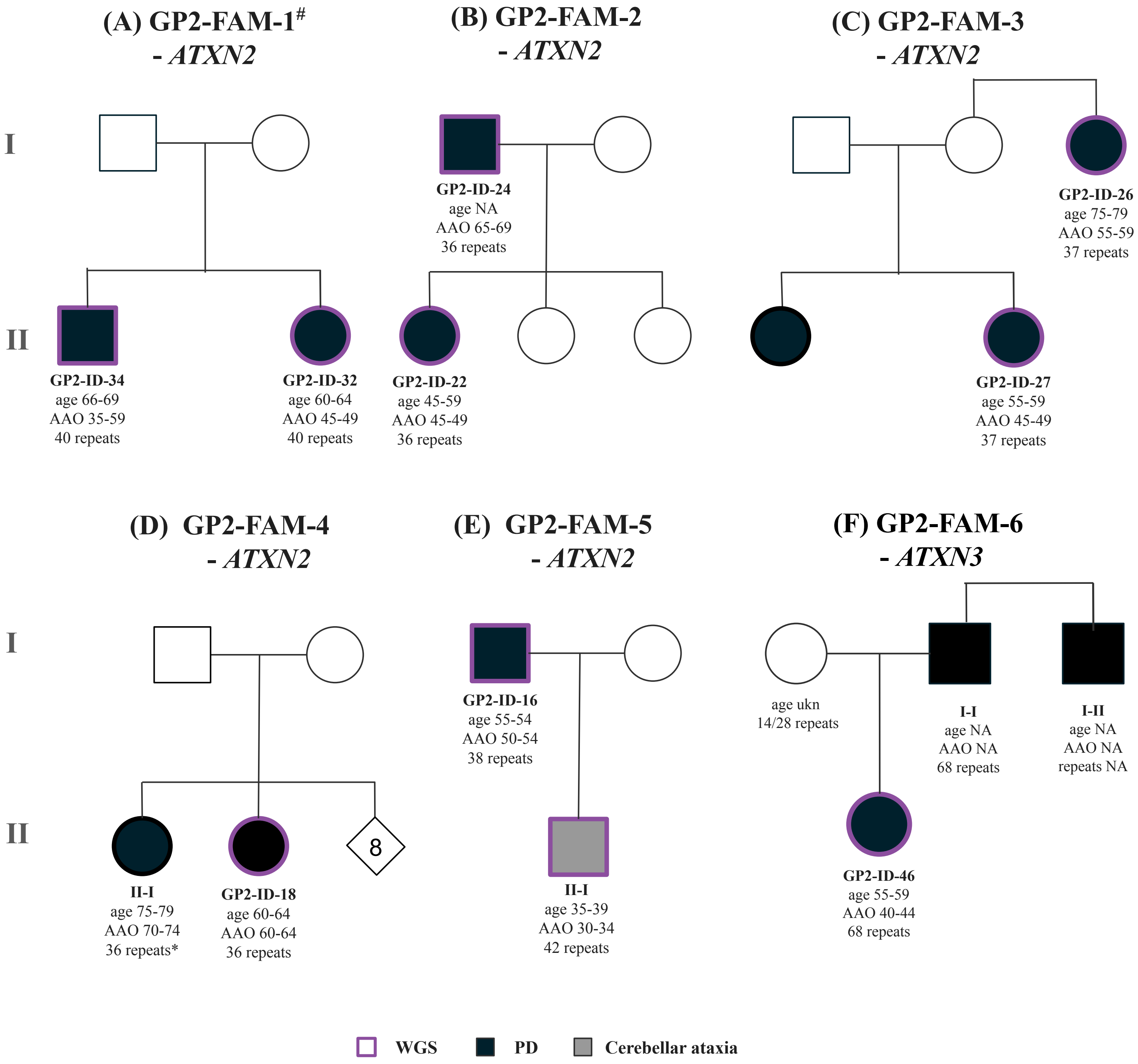


**Supplementary Figure 3. Pedigrees of identified families with multiple affected individuals harboring pathogenic repeat expansions.** The pedigrees were drawn based on kinship analyses and expanded where possible based on reported family history. Affected individuals are indicated by filled symbols: circles (female) and squares (male). Unaffected individuals are indicated by open symbols. Purple circle indicates individuals with genetic data available (WGS). The length of the expanded pathogenic allele is indicated with corresponding age at the sample collection (age) and age at motor symptom onset (if known; AAO) in years.

### Family previously reported by Kim et al. (https://doi.org/10.1016/j.parkreldis.2017.04.003)

* Repeat expansion screening performed outside of this study.

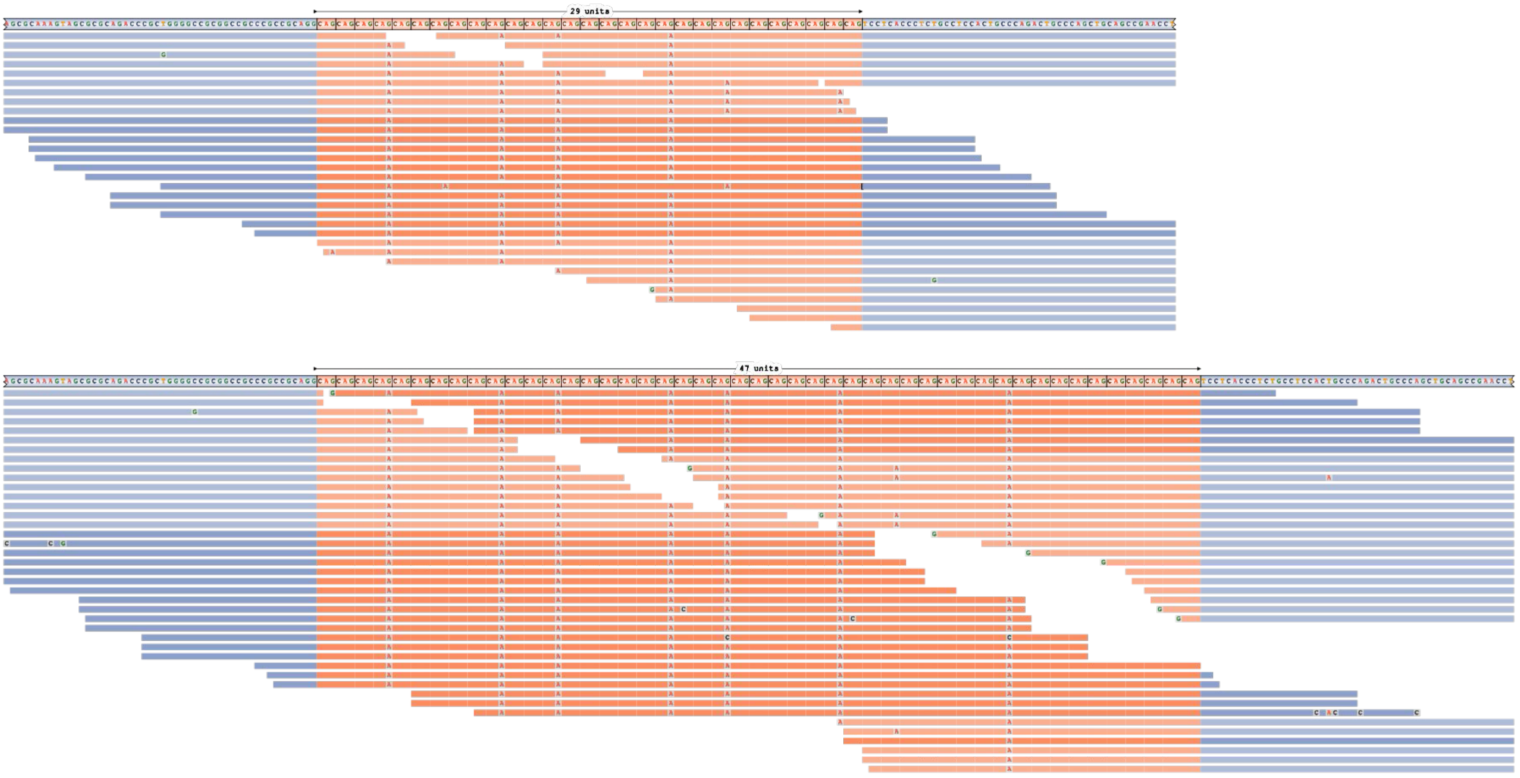


**Supplementary Figure 4 .** ***THAP11*** **pathogenic allele repeat configuration.**

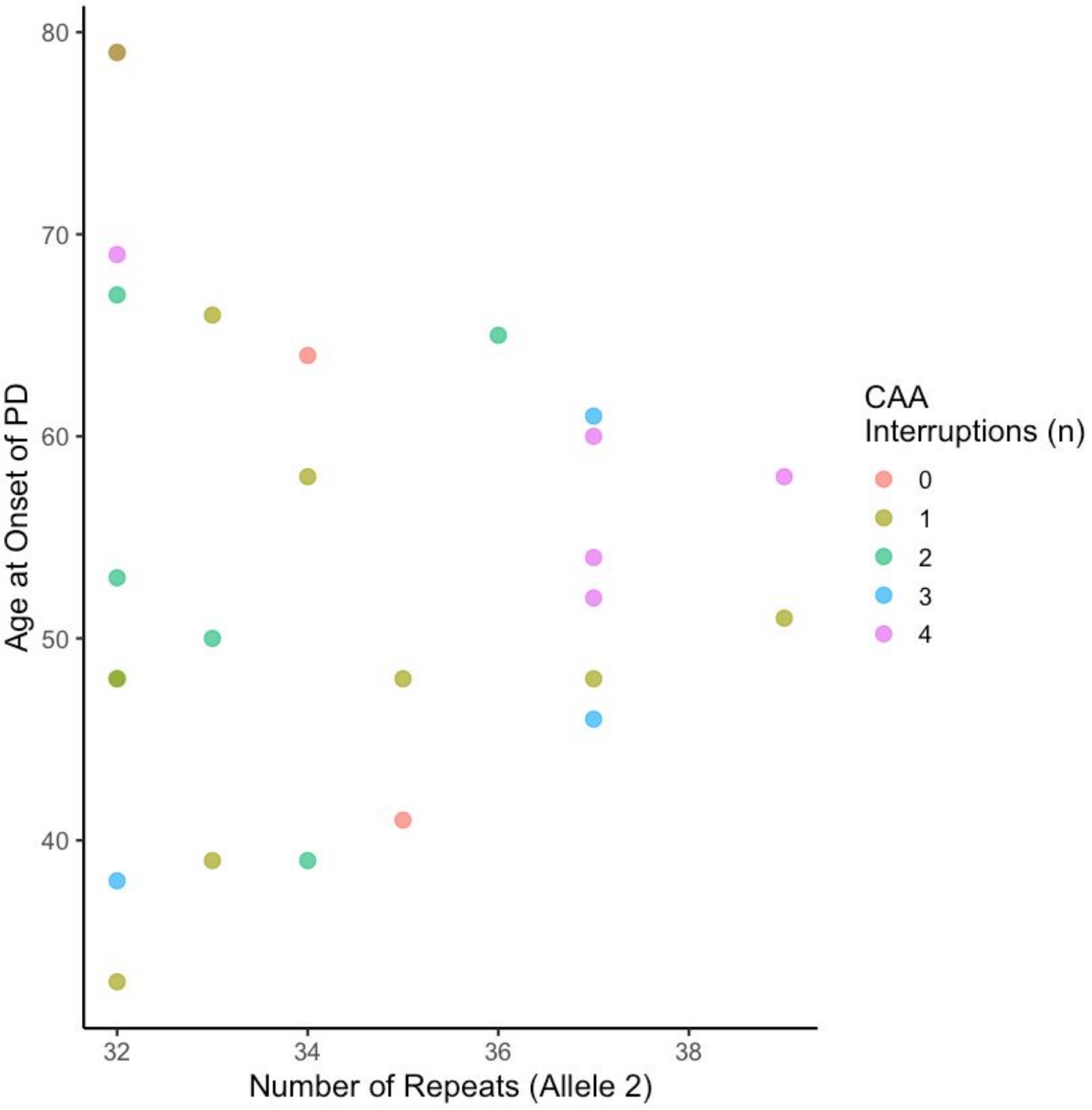


**Supplementary Figure 5. Association of *ATXN2* repeat expansion length (≥32) and CAA interruption count with age at onset in individuals with Parkinson's disease (PD).**
